# Association between frailty assessed by the Clinical Frailty Scale 2.0 and outcomes of acute stroke in older patients

**DOI:** 10.1101/2023.12.05.23299569

**Authors:** Paola Forti, Marianna Ciani, Fabiola Maioli

## Abstract

**Background:** Frailty is a geriatric syndrome characterized by an increased vulnerability to stressors and increased risk of adverse clinical outcomes. While older patients with acute stroke are routinely screened for prestroke disability using the modified Rankin Scale (mRS), because of its known association with stroke outcomes, prestroke frailty is still rarely assessed. The Clinical Frailty Scale (CFS) is a popoular tool for retrospective frailty assessment in the acute setting. The study hypothesis was that prestroke frailty measured with CFS was associated with stroke outcome of older patients independent of prestroke disability assessed with mRS.

**Methods:** We recruited 4086 individuals aged ≥65 years consecutively admitted with acute stroke to an Italian hospital. Prestroke disability (mRS ≥3) was assessed at admission. Prestroke CFS was retrospectively assessed using information from the medical records. Logistic models determined the association of CFS with poor functional outcome, prolonged discharge, unfavorable discharge setting, and poor rehabilitation potential. Cox models determined the association of CFS with 30-day and 1-month mortality. All models were adjusted for prestroke disability and other major confounders.

**Results:** Participants were median age 81 years (25th-75th percentile, 75-87 years), 55.0% female, 82.6% with ischemic stroke, and 26.3% with prestroke disability. Overall prevalence of prestroke frailty (CFS ≥4) was 41.6%. Multivariable-adjusted logistic models showed that CFS was associated with increasing risk of all outcomes except prologed discharge. In severe frailty (CFS 7-8), OR (95%CI) was 3.44 (2.33-5.07) for poor functional outcome, 0.53 (0.38-0.75) for prolonged discharge, 1.89 (0.36-263) for unfavourable discharge, and 6.24 (3.80-10.26) for poor rehabilitation potential (reference CFS 1-3). In multivariable adjusted-Cox models, CFS was unrelated to 30-day mortality but HR (95%CI) of 1-year mortality was significant for both CFS 4-6 (1.70, 1.36-2.11) and CFS 7-8 (1.69, 1.25-2.30).

**Conclusions:** Prestroke frailty measured with CFS was associated with higher risk of several adverse outcomes even after adjustment for prestroke disability and other major confounders.

## Introduction

Frailty is a geriatric syndrome characterized by an increased vulnerability to stressors that is caused by a cumulative decline across multiple physiological systems.^1^ Frailty can explain the heterogeneity in overall health of older persons, it is associated with several adverse clinical outcomes, and its assessment can provide useful prognostic information to guide clinical decision-making and therapeutic interventions in older patients.^2^ Although frail older persons are often disabled and with multiple chronic conditions, frailty is considered a distict entity from both disability and comorbidity.^1^

Stroke is a prototypical stressor event but frailty is rarely mentioned in best practice guidelines on stroke and not yet routinely measured in stroke patients.^3,4^ In current clinical practice and research, prestroke disability retrospectively assessed with the modified Rankin Scale (mRS)^5^ is acknowledged as a robust prognostic predictor of stroke outcomes^6,7^ and, therefore, a commonly used criterion to assist in determining eligibility for hyperacute reperfusion therapies.^8,9^ The mRS is easy to use in a time-pressured setting but was actually designed for assessment of post-stroke functional outcome, with an emphasis on mobility over cognitive disability and comorbidity.^6,9^ Therefore, it may not be a valid measure of pre-stroke global function and overall health, in older persons.^10^ A variety of frailty tools have been developed that widely differ on types of measurements and clinical feasibility.^11^ There is still no standard assessment tool for frailty.^12^ In clinical practice, the choice of a frailty tool depends on the purpose, setting, time availability, and assessor skills.^13^ The Clinical Frailty Scale (CFS)^14^ is one of the most popular tools for retrospective frailty assessment in the acute^15,16^ and hyperacute care setting.^17^ Advantages of CFS include: the tool is based on clinical judgement and does not require performance tests; it focuses on mobility, function, cognition, and comorbidity; it involves a nine-point pictorial scale paired with corresponding text describing classifications of frailty; and it can be readily used by a health care professional without specialist training.^14^ CFS seems a pragmatic and easy choice to assess prestroke frailty in older individuals with acute stroke^4^ but available information about this topic is limited.^18–22^

The primary hypothesis of this study is that prestroke frailty assessed with CFS is associated with stroke outcomes of older patients independent of prestroke disability assessed with mRS. A secondary hypothesis tested in this study is that prestroke CFS and mRS have low agreement and different predictive ability for stroke outcomes.

## Methods

### Study cohort

This is a retrospective observational study based on a single-center cohort of 4094 patients aged ≥65 years who, between January 2006 and December 2018, were consecutively admitted to the Emergency Department (ED) of the Maggiore Hospital (Bologna, Italy) within 24 hours after onset of acute stroke and subsequently transferred to the local Stroke Unit (SU). All patients had at least one CT-head scan at ED admission. Only patients with a final diagnosis of ischemic stroke (IS) and primary intracerebral hemorrhage (ICH) were considered eligible. Patients with transient ischemic attack, subarachnoid hemorrhage, and secondary ICH due to underlying vascular malformations and intracranial tumors were excluded. Stroke severity at admission was assessed using the National Institutes of Health Stroke Scale (NIHSS) score,^23^ categorized as <6, 6-15, and >15.^24^ Patients were treated according to standard guidelines for management of acute stroke.^25^ In IS patients, the decision to perform reperfusion therapies (intravenous thrombolysis using the fibrinolytic agent alteplase or endovascular thrombectomy) was based on clinical judgment.

### Prestroke frailty and disability

At the time of ED admission, the stroke consultant on duty retrospectively assessed mRS in the two preceeding weeks (pre-mRS) based on an unstructured direct interview with the patient and, whenever possible, a knowledgeable proxy (relatives and usual caretakers). Prestroke disability was defined as pre-mRS ≥3.^26^ Concurrent frailty level according to CFS version 2.0^27^ (**Figure 1**) was retrospectively assessed by two authors (P.F. and M.C.) based on any available information on premorbid mobility, function, cognition, and medical history as documented in the patient chart. Prestroke frailty was defined as CFS ≥ 4 and further graded as mild-to-moderate (CFS 4 to 6) and severe-to-terminal (CFS 7 to 8).^27^ Patients with CFS 9 were excluded from further analyses because this level identifies persons whose life expectancy is shorter than six months but are not otherwise living with frailty, which disrupts the progression of frailty from CFS 1 to 8.^28^ Similarly to what would happen in real clinical practice, CFS raters were not blind to pre-mRS. Retrospective CFS scoring using information not primarily intended for this goal has been validated with good reliability and accuracy.^29,30^

**Figure 1.**
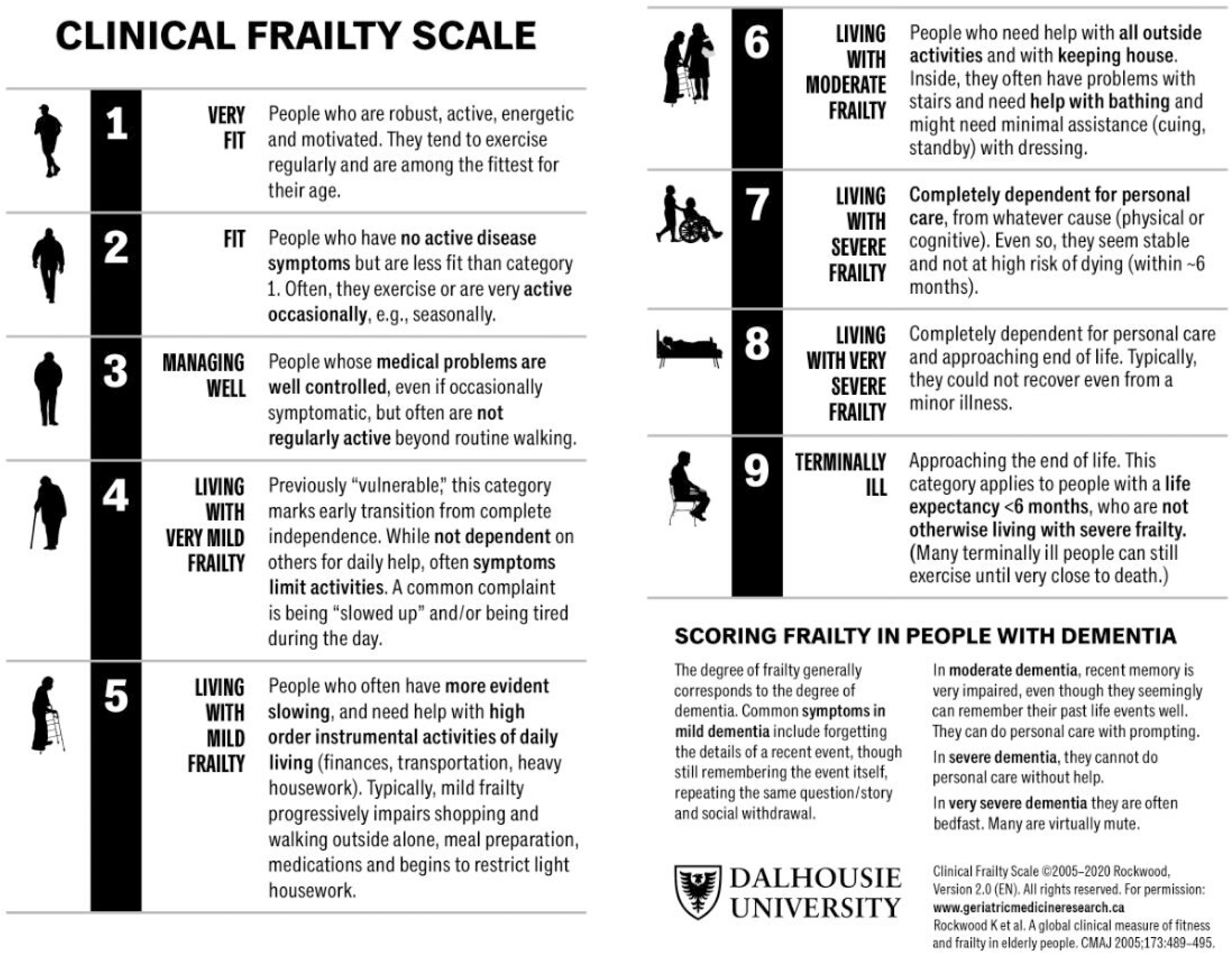
The Clinical Frailty Scale version 2.0. Printed with permission from copyright holder.

### Other prestroke variables

Baseline demographic and clinical data were extracted from the patient charts. Comorbidity was defined using the Charlson Comobidity Index (CCI)^31^, which takes into account the number and severity of 19 pre-defined comorbid conditions. CCI score was categorized as 0, 1, 2-3, and ≥4.^32^

### Outcomes

SU attending physicians re-assessed mRS at discharge (discharge-mRS). Poor functional outcome was defined as discharge-mRS >1 for patients with pre-stroke mRS <2 and as discharge-mRS greater than pre-mRS for patients with pre-mRS 2-4; patients with pre-mRS 5 were excluded because they would have a favourable outcome just surviving.^10^ Prolonged discharge was defined as length of SU stay >7 days.^33^ Discharge destination was defined “favorable” (home or rehabilitation facility) or “unfavourable” (long-term care facility, another acute hospital setting, or in-hospital death).^34^ In a subgroup analysis, poor rehabilitation potential, defined as discharge to long-term care, was contrasted to discharge to rehabilitation facilities. All-cause mortality at 30-day and 1-year after stroke onset was ascertained from the Italian Regional Mortality Registry. In IS patients treated with reperfusion therapies, percentage improvement in NIHSS after treatment was calculated as (admission NIHSS score minus discharge NIHSS score)×100/admission NIHSS score.

### Statistical analysis

Data are reported as count (percentages) for categorical variables or median (25^th^-75^th^ percentile) for continuous variables. Univariate comparisons between groups were performed using chi-square test or Kruskal-Wallis test as appropriate. Agreement between CFS and pre-mRS was assessed using weighted kappa^35^. Predictive ability of both scales for the study outcomes was estimated using the Area under the Receiver Operating Characteristic curves (AuROC). Confidence intervals were calculated according to binomial exact formula. DeLong’s method was used for AuROC comparison.^36^

The association of CFS categories with all study outcomes excepting mortality was assessed using Odds Ratios (OR) and their 95% confidence intervals (95%CI) from logistic models. The association of CFS categories with mortality was assessed using Hazard Ratios (HR) and their corresponding 95%CI from Cox proportional hazard models. A priori chosen confounders for adjusted models included: age, sex, CCI score, stroke subtype, admission NIHSS, and prestroke disability. Effect modification of CFS by model confounders was systematically assessed and interactions were considered significant for p-value < 0.010. In subanalyses of treated IS patients, confounders for adjusted models included age, sex, CCI score and prestroke disability. The association of CFS categories with NIHSS improvement was assessed using linear regression. For the other study outcomes, treated IS patients were classified as frail vs non-frail because of insufficient sample size. Analyses were performed with R software version 3.5.3. Significance for P value was set at the 0.050 (two-tailed).

### Standard Protocol Approvals, Registrations, and Patient Consents

At SU admission, written informed consent for research use of their medical records was sought from patients or their legally authorized representatives according to the Declaration of Helsinki. The Maggiore Hospital Ethics Committee approved the study (approval number CE16092).

### Data Availability

Data for this study will be made available by request from any qualified investigator.

## Results

### Clinical and demographic factors

After exclusion of eight persons with CFS 9, the final cohort included 4086 patients; 2249 were women (55.0%) and median age was 81 years (25th-75th percentile, 75-87 years). Overall prevalence of prestroke frailty was 41.6% (29.4% for CFS 4-6 and 12.2% for CFS 7-8). Only twelve patients were classified as CFS 1 (“very fit”). ***Supplemental Figure F1*** shows how frailty prevalence increased from 14.7% at age 65-69 to 78.7% at age ≥90 years.

**Table 1** shows how increasing CFS was associated with age, female sex, living in nursing home before admission, prestroke disability, and comorbidity. IS was the most frequent stroke subtype in all CFS categories and stroke severity increased with increasing fraily. Occurrence of all study outcomes increased across increasing CFS categories excepting prolonged discharge, which was lowest for CFS 7-8. ***Supplemental Table T1*** provides information about the number of patients for each outcome by CFS category. ***Supplemental Figure F2*** provides information about the distribution of pre-and discharge-mRS across CFS categories.

**Table 1.**
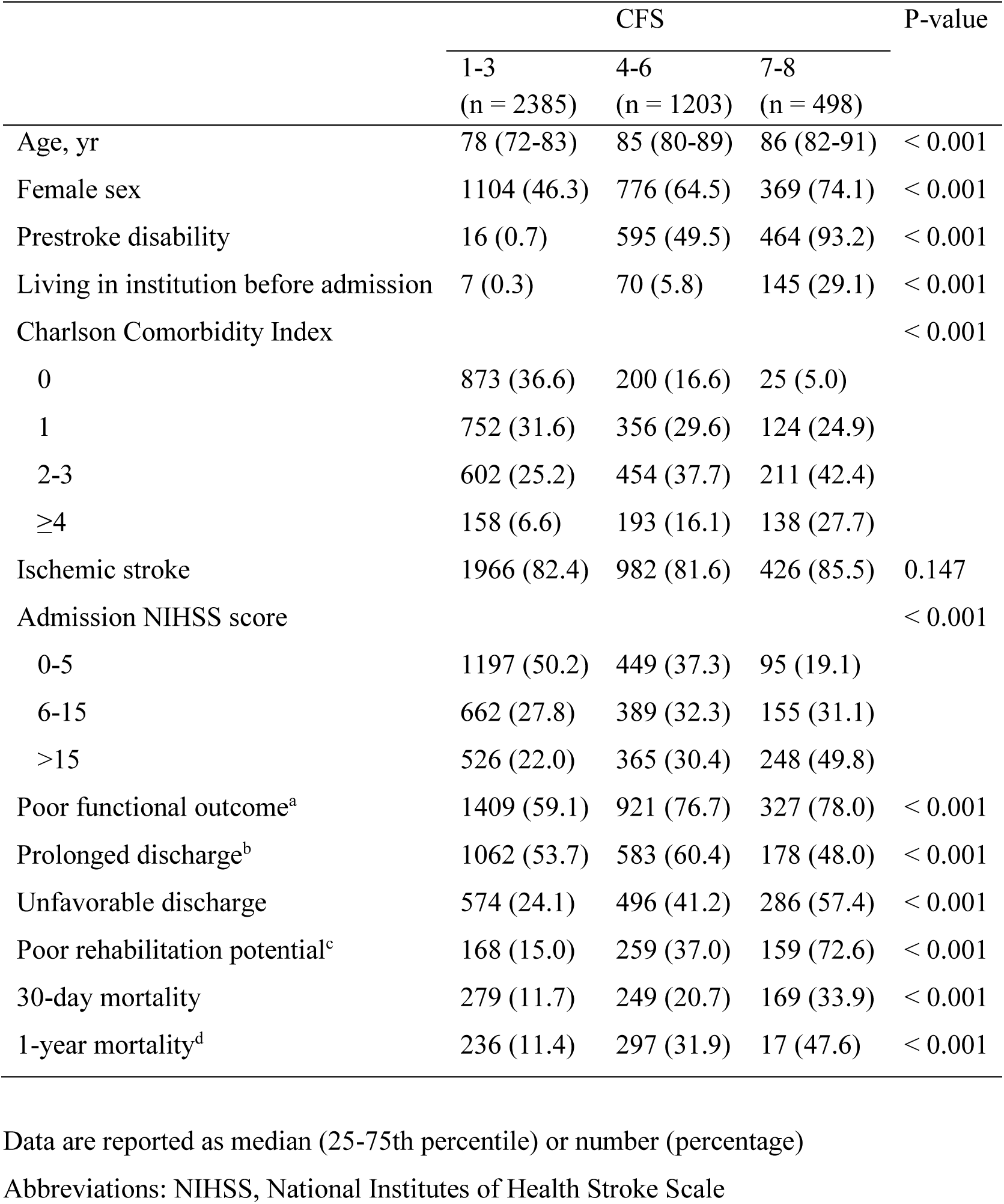
Population characteristics by Clinical Frailty Score (CFS)

|  | CFS |  |  | P-value |
| --- | --- | --- | --- | --- |
|  | 1-3<br>(n = 2385) | 4-6<br>(n = 1203) | 7-8<br>(n = 498) |  |
| Age, yr | 78 (72-83) | 85 (80-89) | 86 (82-91) | < 0.001 |
| Female sex | 1104 (46.3) | 776 (64.5) | 369 (74.1) | < 0.001 |
| Prestroke disability | 16 (0.7) | 595 (49.5) | 464 (93.2) | < 0.001 |
| Living in institution before admission | 7 (0.3) | 70 (5.8) | 145 (29.1) | < 0.001 |
| Charlson Comorbidity Index |  |  |  | < 0.001 |
| 0 | 873 (36.6) | 200 (16.6) | 25 (5.0) |  |
| 1 | 752 (31.6) | 356 (29.6) | 124 (24.9) |  |
| 2-3 | 602 (25.2) | 454 (37.7) | 211 (42.4) |  |
| ≥4 | 158 (6.6) | 193 (16.1) | 138 (27.7) |  |
| Ischemic stroke | 1966 (82.4) | 982 (81.6) | 426 (85.5) | 0.147 |
| Admission NIHSS score |  |  |  | < 0.001 |
| 0-5 | 1197 (50.2) | 449 (37.3) | 95 (19.1) |  |
| 6-15 | 662 (27.8) | 389 (32.3) | 155 (31.1) |  |
| >15 | 526 (22.0) | 365 (30.4) | 248 (49.8) |  |
| Poor functional outcome<sup>a</sup> | 1409 (59.1) | 921 (76.7) | 327 (78.0) | < 0.001 |
| Prolonged discharge<sup>b</sup> | 1062 (53.7) | 583 (60.4) | 178 (48.0) | < 0.001 |
| Unfavorable discharge | 574 (24.1) | 496 (41.2) | 286 (57.4) | < 0.001 |
| Poor rehabilitation potential<sup>c</sup> | 168 (15.0) | 259 (37.0) | 159 (72.6) | < 0.001 |
| 30-day mortality | 279 (11.7) | 249 (20.7) | 169 (33.9) | < 0.001 |
| 1-year mortality<sup>d</sup> | 236 (11.4) | 297 (31.9) | 17 (47.6) | < 0.001 |

Data are reported as median (25-75th percentile) or number (percentage)
Abbreviations: NIHSS, National Institutes of Health Stroke Scale

### Agreement between CFS and pre-mRS

Overall prevalence of prestroke disability was 26.3%. Prestroke frailty was identified in 21.3% of those without prestroke disability. Prestroke disability was recorded only in 0.7% of non-frail patients. **Figure 2** shows how pre-mRS levels 0 to 2 and CFS levels 1 to 3 tended overlap while there was a noticeable dispersion of pre-mRS levels across CSF levels above 3, even with some patients classified as totally independent according to pre-mRS but moderately to severely frail according to CFS. Weighted kappa for CFS and pre-mRS was 0.56, suggestive of only moderate agreement.^35^

**Figure 2.**
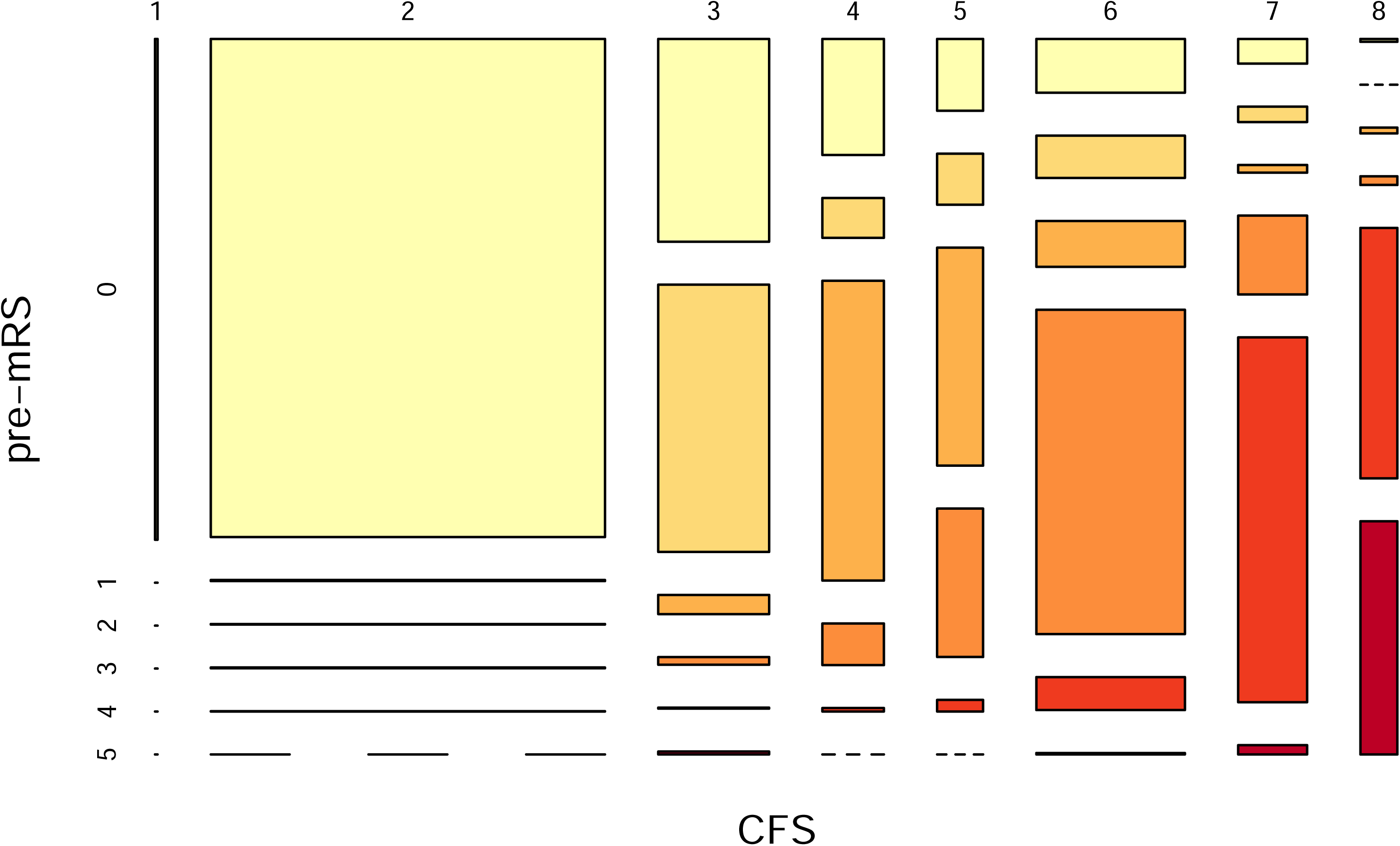
Mosaic plot representing the study patients stratified by levels of Clinical Frailty Scale (CFS, horizontal axis) and prestroke modified Rankin Scale (pre-mRS, vertical axis). The width of the colums represents the number of observation for each CFS level. The height of each bar represent the number of observations for individual pre-mRS levels within each CFS level.

### Predictive ability of CFS and pre-mRS for the study outcomes

Comparison of AuROCs (**Table 2**) showed that CFS was statistically superior to pre-mRS for prediction of poor functional outcome, unfavorable discharge, and poor rehabilitation potential while no difference was found for the other study outcomes. However, AuROCs for both scales were below the traditional 0.80 threshold for clinical usefulness.^37^

**Table 2.** Comparison of Clinical Frailty Scale (CFS) and preadmission modified Rankin scale (pre-mRS) for prediction of the study outcomes.

|  | **CFS** | **pre-mRS** | **P-value** |
| --- | --- | --- | --- |
| Poor functional outcome | 0.605 (0.588-0.621)* | 0.581 (0.564-0.597) | < 0.001 |
| Prolonged discharge | 0.502 (0.483 – 0.520) | 0.506 (0.488 – 0.524) | 0.289 |
| Unfavorable discharge | 0.648 (0.630-0.666) | 0.635 (0.618-0.652) | 0.003 |
| Poor rehabilitation potential | 0.737 (0.713-0.761) | 0.721 (0.697-0.745) | 0.013 |
| 30-day mortality | 0.639 (0.617-0.662) | 0.637 (0.616-0.659) | 0.710 |
| 1-year mortality | 0.703 (0.681-0.725) | 0.700 (0.678-0.723) | 0.537 |

Data are areas under the curve (95% Confidence Intervals). P-values are for non-parametric comparison of CFS and pre-mRS estimates.

### Multivariable-adjusted association of CFS with the study outcomes

Figure 3 summarizes the results from adjusted logistic (Panel A) and Cox models for the association of CFS with the study outcomes. Likelihood of poor functional outcome, unfavorable discharge, and poor rehabilitation potential increased with increasing frailty level while likelihood of prolonged discharge was almost halved for CFS 7-8 compared to CFS 1-3. Results for unfavorable discharge did not change when excluding 222 patients already living in institution before hospital admission (CFS 4-6: adjusted-OR, 1.46, 95%CI 1.17-1.82; CFS 7-8: adjusted-OR 2.66, 95%CI 1.85-3.81). There was no significant association of CFS with 30-day mortality but 1-year mortality risk was significantly increased for both CFS 4-6 and 7-8 compared to CFS 1-3. In all models, both the independent effect of prestroke disability on outcome prediction and its interaction with CFS categories were not significant (p-value > 0.200 for all).

**Figure 3.**
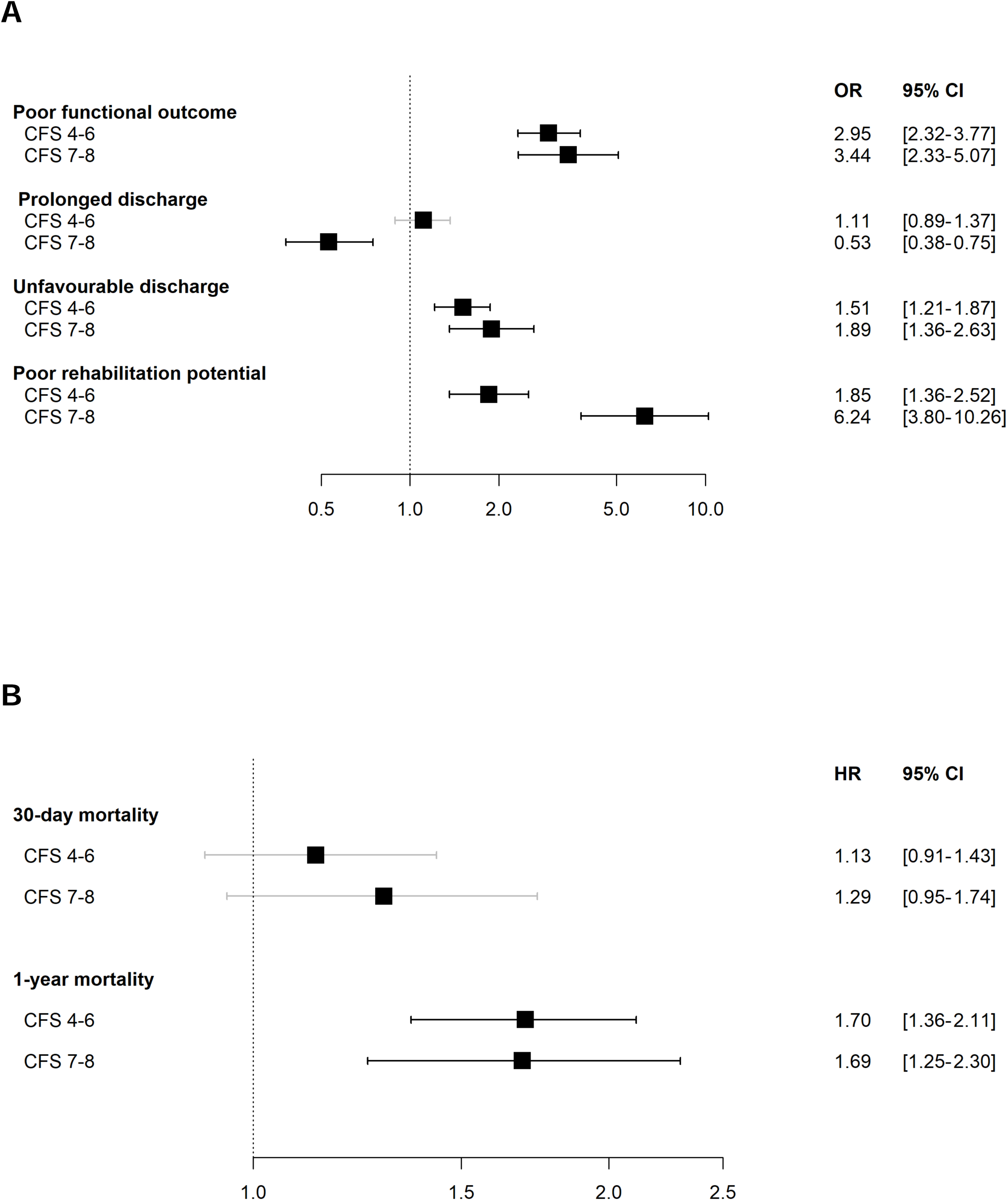
Multivariable-adjusted associations of Clinical Frailty Scale (CFS) with the study outcomes from multivariable-adjusted models. CFS was categorized as no frailty (score 1-3), mild to moderate frailty (score 4-6) and severe frailty (score 7-8). Panel A shows odds ratios (OR) and their 95% confidence intervals (95% CI); panel B, hazard ratios (HR) and their 95% CI. All models included age, sex, Charlson Comorbidity Index score, stroke subtype, admission NIHSS, and prestroke disability.

### IS patients treated with reperfusion therapies

There were 3374 patients with IS (82.6%). Cardioembolism was the most frequent etiology (35.2%), followed by cryptogenic (33.4%), small vessel occlusion (19.9%), large artery atherosclerosis (9.9%), multiple causes (1.0%) and other causes (0.6%). Reperfusion therapies were performed in 498 patients (14.8%): 83.5% received intravenous thrombolysis, 4.2% mechanical thrombectomy, and 12.2% both. Only 25.9% of treated patients were frail compared to 44.5% of non-treated patients but adjusted-OR was not significant (1.00, 95%CI 0.74-1.34). Among treated patients, overall prevalence of prestroke disability was only 12% but the proportion was much higher in frail (45.7%) compared to non-frail patients (just one case, 0.3%). Figure 4 shows the results from adjusted logistic (Panel A) and Cox models (Panel B) for the association of frailty with the study outcomes in treated IS patients. Significant associations were found for poor functional outcome (74% of 127 frail vs 43.6% of 369 non-frail patients) and poor rehabilitation potential (40.9% of 66 frail vs 8.6% of 139 non-frail patients) but not for prolonged (42.3% of 97 frail vs 34.9 of 278 non-frail patients) and unfavorable discharge (45.4% of 129 frail vs 27.9% of 369 non-frail patients). Because of the small number of cases (n=77), only the association of frailty with overall mortality during a 1-year follow-up could be estimated. Mortality rate was higher in frail compared to non-frail patients (36.1% vs 8.4%) and the corresponding adjusted-HR was 2.36, 95%CI 1.24-4.50.

**Figure 4.**
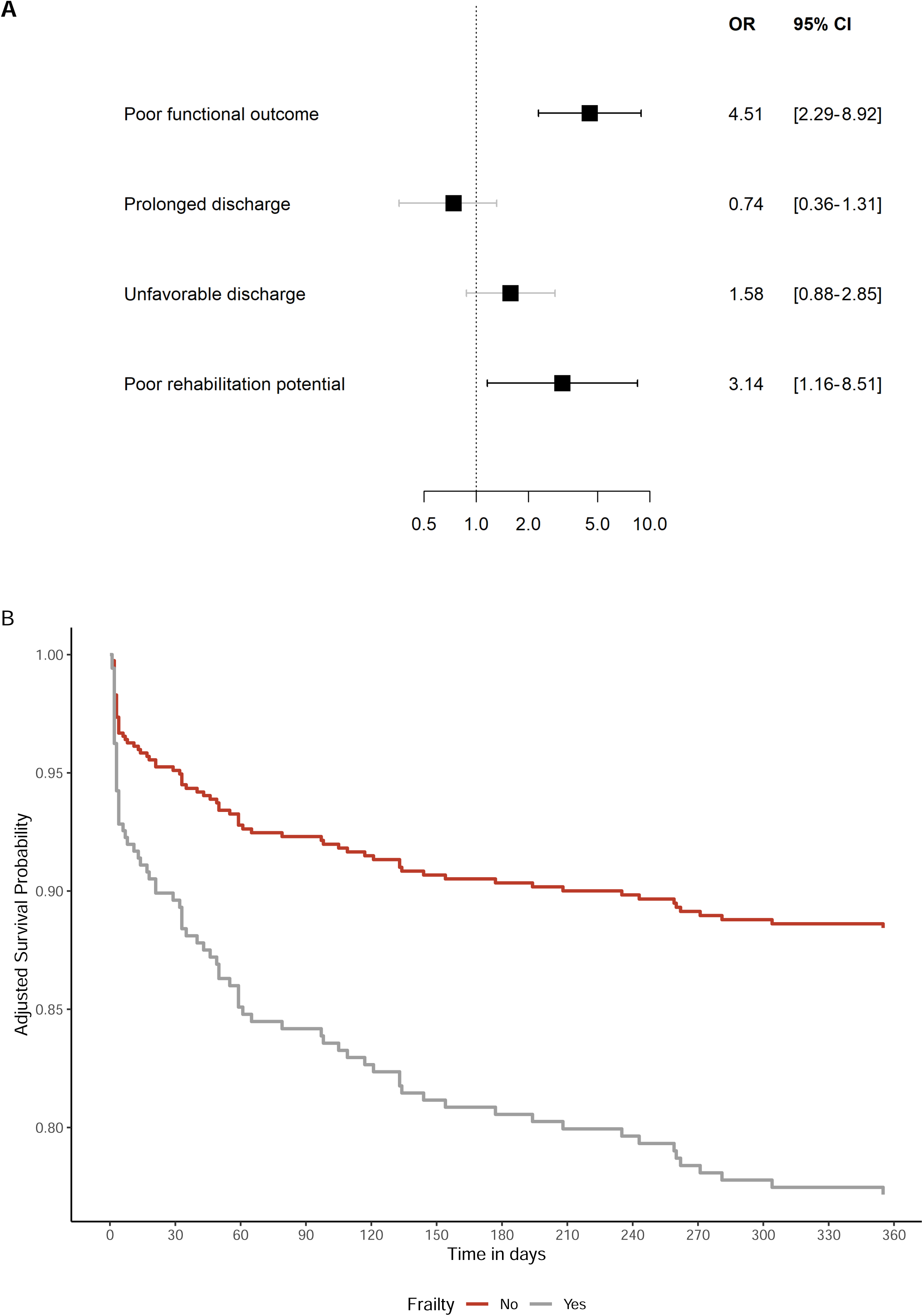
Multivariable-adjusted associations of frailty with the study outcomes in patients with ischemic stroke treated with reperfusion therapies. Frailty was defined as Clinical Frailty Scale ≥4. Panel A shows odds ratios (OR) and their 95% confidence intervals (95% CI). Panel B shows the adjusted survival probability curves for frail and non-frail patients. All models included age, sex, Charlson Comorbidity Index score, admission NIHSS, and prestroke disability.

Among treated patients, stroke severity was higher in frail than non-frail persons (31.0% vs 19% with admission NIHSS >15, p-value = 0.012). However, median percentage improvement in NIHSS after reperfusion therapy had a U-shaped relationship with CFS score (***Supplemental Figure F3***). The adjusted linear model confirmed that median NIHSS improvement for CFS 1-3 (71%, 35%-100%) was higher compared to CFS 4-6 (59%, 12%-79%, p-value = 0.003) but did not differ compared to CFS 7-8 (78%, 49-84%, p-value = 0.708).

In all of the above reported models, both the independent effect of prestroke disability on outcome prediction and its interaction with frailty were not significant (p-value > 0.200 for all).

## Discussion

This cohort study of older patients with acute stroke showed that prestroke frailty retrospectively assessed using CFS version 2.0 was highly prevalent and may convey additional prognostic information for several stroke outcomes with respect to prestroke disability assessed using mRS.

Previous studies of frailty occurrence in older stroke patients produced inconsistent results, mainly because of the great heterogeneity in the operational definition of frailty (estimates ranging from 2% to 68%).^3,38^ Most of these studies are not comparable with ours because they used variations of the physical frailty phenotype, which is based on motor and activity measures, or frailty cumulative indexes, which variably incorporate measures of function, cognition, comorbidities, and social factors.^2^ Two studies of IS patients using CFS reported frailty prevalence estimates ranging between 38%^20^ and 54%,^18^ which agree with our findings. However, comparability is limited because these studies used an older version of CFS, in which level 5 is the first including the term frail in its description^14^ and, accordingly, frailty was defined as CFS ≥5. The association of CFS with age, female gender, and comorbidity found in our cohort is not surprising because it reflects known general features of frailty.^2^ In this older cohort, we also found no difference in frailty prevalence by stroke type. A metanalysis of 24 studies reported that frailty was more prevalent in IS than ICH,^38^ but this result may derive from the inclusion of studies involving patients aged less that 65 years, with data for ICH mostly coming from younger cohorts.

Our investigation provided evidence of unfavourable associations between increasing CFS and all of the study outcomes excepting prolonged discharge. The associations of CFS with outcomes related to discharge setting, functional status at SU discharge, and mortality can have several explanations: acute stroke and frailty can exacerbate each other and trigger a self-propagating cycle; frailty can also influence several physical and non-physical aspects of stroke recovery, including acute complications (in particular delirium and infections), malnutrition, post-stroke cognitive impairment, and effectiveness of rehabilitation and psychosocial interventions.^4,39^ Our frailest patients also had a shorter SU stay, but our study design does not allow to understand whether this happened because frailest patients were correctly spared futile diagnostic and therapeutic procedures or actually received suboptimal care.

Existing evidence that frailty can provide prognostic information additional to major stroke predictors such as age, stroke severity, and pre-mRS is still weak. A metanalysis of 14 stroke studies^3^ reported associations of frailty with longer hospital stay, poor functional outcome, discharge destination, and long-term mortality. However, the authors stressed the low number of eligible studies for the individual outcomes and the heterogeneity of the tools employed for frailty identification (only one study used CFS^18^).

Available information about the agreement of prestroke frailty measures with pre-mRS is also scant and their correlation appears to be, at the best, only moderate, with no clear dose-response patterns.^3,4^

There are only four previous studies of CFS in older stroke patients admitted to SU, all of them using the older version of CFS scale.^14^ In a study of 433 IS patients aged ≥75 years,^18^ CFS had no significant correlation with pre-mRS; it was associated with 28-day mortality in the main cohort but unrelated to NIHSS improvement after 24 hours in 63 patients undergoing reperfusion therapy. Another study^20^ investigated 472 IS patients aged ≥65 years admitted to a stroke center able to provide only intravenous thrombolysis (patients with CFS 9 were excluded). CFS was weakly correlated with pre-mRS and predicted 28-day and 1-yr mortality but did not predict either discharge setting in the main cohort and NIHSS improvement in 178 patients undergoing thrombolisis. AuROC comparisons of CFS and pre-mRS showed that CFS offered a slight predictive advantage for mortality but not for discharge setting. Similarly to our results, all AuROC estimates were below the 0.80 threshold. Noticeably, both the above mentioned studies tested that the association of CFS with stroke outcomes was independent of age, stroke severity and comorbidities but did not include pre-mRS as a confounder.

Two other studies using CFS focused on select subsets of IS older patients without prestroke disability who underwent mechanical thrombectomy. In 159 patients aged ≥80 years, frailty was associated with poor functional outcome at 3 months (mRS >3) and 1-year mortality.^22^ In 198 patients aged ≥70 years, there was low agreement between CFS and pre-mRS and frailty (here defined as CFS ≥ 4) was associated with mRS at 90-day but not with length of hospital stay.^21^

In our stroke cohort, AuROC curves showed that CFS had a slightly better prognostic ability than pre-mRS for some study outcomes but neither scale attained clinical meaningfulness for any. However, agreement between the scales was low and multivariable analyses confirmed that CFS provided prognostic information independent of prestroke disability measured with mRS. Additionally, the study showed that prestroke frailty, independent of prestroke disability, was associated with several adverse outcomes of IS patients treated with reperfusion therapies. This agrees with results from a metanalysis of nine studies^40^ (including all the above mentioned four studies using CFS) that reported an association of frailty with poor functional outcome and 1-year, but not 1-month, mortality in IS treated patients.

The U-shaped association we found between CFS and NIHSS improvement after reperfusion therapies is particularly interesting. While it can be reasonable to expect that persons with mild-to-moderate frailty are more susceptible to acute ischemic damage, the potential for neurologic improvement in patients with severe frailty may appear counterintuitive. However, several, not mutually exclusive explanations can be proposed. First, in patients with severe frailty, pre-existent motor and cognitive impairment may interfere with a reliable estimation of neurological impairment on hospital admission,^39^ especially when using NIHSS that is not intended to distinguish prestroke and acute deficits.^23^ Second, patients with severe frailty might present with greater neurological severity at IS onset independent of their actual structural ischemic damage because of a lower brain reserve, concurrent confusional states, and comorbidities adding their confounding effects.^41–43^

There is a strong evidence that patients with pre-stroke disability have a higher risk of unfavourable stroke outcomes and may derive less benefit from hyperacute reperfusion therapies.^6,7^. However, mRS was not designed to measure prestroke disability; the wording of its grades in not suited to pre-stroke assessment; and the tool may misestimate a patient’s ability to recover because it overvalues physical disability with respect to cognition and general health.^6,9^ At present, both European^8^ and US^9^ stroke guidelines on reperfusion therapies acknowledge that exclusion of patients on the sole ground of prestroke disability may not be justified.

The main strength of this study is the use of CFS version 2.0 for frailty identification in a large elderly cohort taken from a a “real world” pool of consecutive, unselected stroke admissions.

This study has also several limitations. First, the retrospective design does not allow for establishing causal links. Second, a single-center cohort limits generalizability of results. Third, pre-mRS scoring occurred at the time of ED setting, while evaluating whether the patient was eligible for reperfusion therapy, and it was performed by multiple observers. By contrast, CFS was scored post hoc, by two raters with similar background who were spared the time pressure often occurring during hyperacute management of stroke.

Moreover, CFS scoring was based on information from the whole medical chart and some data may have not been available when pre-mRS was scored. A fourth limitation is that, since analyses were based on observational data, we had to adjust for baseline differences and many important confounders may have been left out. The relatively small size of the IS treated subset is another drawback that limits generalization of the study findings. Moreover, as reperfusion treatment decisions for patients with prestroke disability were not standardized, no conclusion can be drawn about the efficacy and safety of these procedures in our cohort. However, as the use of reperfusion therapy in elderly persons with prestroke disability is still generally low,^8,9^ we believe that our data may be of interest and promote further investigations. A final limitation is the lack of information on death causes and post-discharge functional outcome. Moreover, we used discharge destination as a surrogate measure of post-stroke function and rehabilitation potential. Although we ascertained that results for these outcomes did not change after exclusion of patients with pre-existing disability already living in nursing homes prior to admision, the potential for misclassification must be acknowledged.

In conclusion, our data suggest that CFS and pre-mRS are not a substitute for each other and CFS may help to inform decisions in elderly patients admitted with acute stroke. CFS has several advantages over other frailty tools: it encompasses a broad assessment of frailty based on clinical judgement, is quick, and does not rely on direct measurement of specific items, specialized equipment, or extra staff.^4^ Further work exploring the relevant prognostic properties of CFS in acute stroke patients is necessary, particularly with respect to treatment decisions.

## Supplemental Material

Tables T1

Figures F1-F3

